# A flexible pipeline for reproducible exome-wide rare variant gene-trait associations

**DOI:** 10.1101/2025.06.26.25330270

**Authors:** Kevin Y. H. Liang, Ethan Kreuzer, Yann Ilboudo, Yiheng Chen, J. Brent Richards, Guillaume Butler-Laporte

## Abstract

**Summary:** Exome-wide gene-burden association studies are widely used to assess gene – trait relationships. By focusing on coding variants, such analyses can directly quantify the magnitude and direction of a gene’s effect on a given trait across the proteome, information that cannot be easily derived from genome-wide associate studies. However, the lack of a standardized workflow poses a significant challenge for reproducibility. Here, we provide a customizable workflow implemented in Python 3 and Nextflow for performing exome-wide rare variant gene – trait association testing. We demonstrated its utility by replicating three recent studies. This workflow will also serve as a framework for performing similar analyses in a standardized and systematic manner.

**Availability and Implementation:** This workflow is publicly available at https://github.com/richardslab/EXWAS_pipeline under the MIT license.

**Supplementary information:** Supplementary information will be made available online.

## Introduction

Exome-wide association studies (ExWAS) are widely used to assess gene-trait associations^1,2^. The focus on coding variants allows ExWAS to directly estimate the association of genetic variants attributable to a specific gene with a trait, which cannot be easily derived from genome-wide association studies (GWAS), as these generally find non-coding associations. Consequently, ExWAS has validated several known disease drug targets in epileptic encephalopathy^3^, low-density cholesterol^2^, body-mass index (BMI)^4^, and osteoporosis^5^. Additionally, it has revealed novel gene-trait associations such as the effect of the *BSN* gene on BMI^4^, the *INHBE* gene on abdominal obesity^6^, and the *CD109* gene on bone mineral density^5^ – to name a few.

One important limitation of ExWAS is reduced statistical power as pathogenic coding variants tend to be rare due to strong natural selective pressure^7^. Consequently, much larger sample sizes are required to have a sufficient number of carriers. A common method to partially overcome this limitation, and the focus of this study, is gene burden tests (or gene collapsing tests) where qualifying variants of a gene are collapsed into a single metric.

ExWAS gene burden tests have two important parameters to consider. First is the definition of qualifying variants. Several algorithms are available to predict variant consequences^8^. Importantly, the definition of qualifying variant significantly affects association results^9^. Second, it is important to consider different gene-burden test statistics as each is based on distinct biological assumptions. For instance, the standard additive effect gene burden test is optimal when all qualifying variants influence the trait in the same direction. However, if qualifying variants have either protective or deleterious effects, SKAT or SKAT-O are more suitable^9^. Importantly, statistical power can vary considerably based on the underlying biological assumptions, which are not known in advance^10^. Consequently, it is important to evaluate different combinations of variant inclusion criteria and burden test statistics to draw robust conclusions from ExWAS. Moreover, replication in independent cohorts is recommended whenever possible^10^.

Currently, there are no standardized workflows for performing ExWAS gene burden tests which poses an important challenge for replication and comparison. Here, we present a customizable workflow for performing ExWAS gene burden tests and variant annotations on biobank scale data using well-established bioinformatics tools. This workflow is written in Python and Nextflow ensuring portability and compatibility across different computing environments. It is actively managed with regular updates and the addition of new analytical workflows. This tool is publicly accessible on GitHub (https://github.com/richardslab/EXWAS_pipeline).

## Design and implementation

### Pipeline overview

The objective of this pipeline is to provide standardized workflows for running gene burden tests using standard input files. This pipeline is implemented in Python 3 and Nextflow and uses well-established tools for data processing and association testing. Specifically, it relies heavily on BCFtools and Plink v1.9 for manipulating genetic data, VEP for variant annotation (with plugins, as described below), and Regenie for association testing.

This pipeline is divided into 4 workflows. The first is to containerize VEP within an Apptainer^11^ image based on user specification. This simply executes the Apptainer build command with user inputs and will not be discussed further. Apptainer is a an open-source container platform which can run securely on most computing clusters (specifically, it can run without the need to give administrator privilege to users). The remaining 3 workflows are described in more details below. The outputs of all four workflows are outlined in Figure 1 and Table S1. Runtime parameters are specified by two configuration files. Configuration file formats are described on the GitHub repository (https://github.com/richardslab/EXWAS_pipeline/) along with file templates and will not be discussed here.

**Fig 1:**
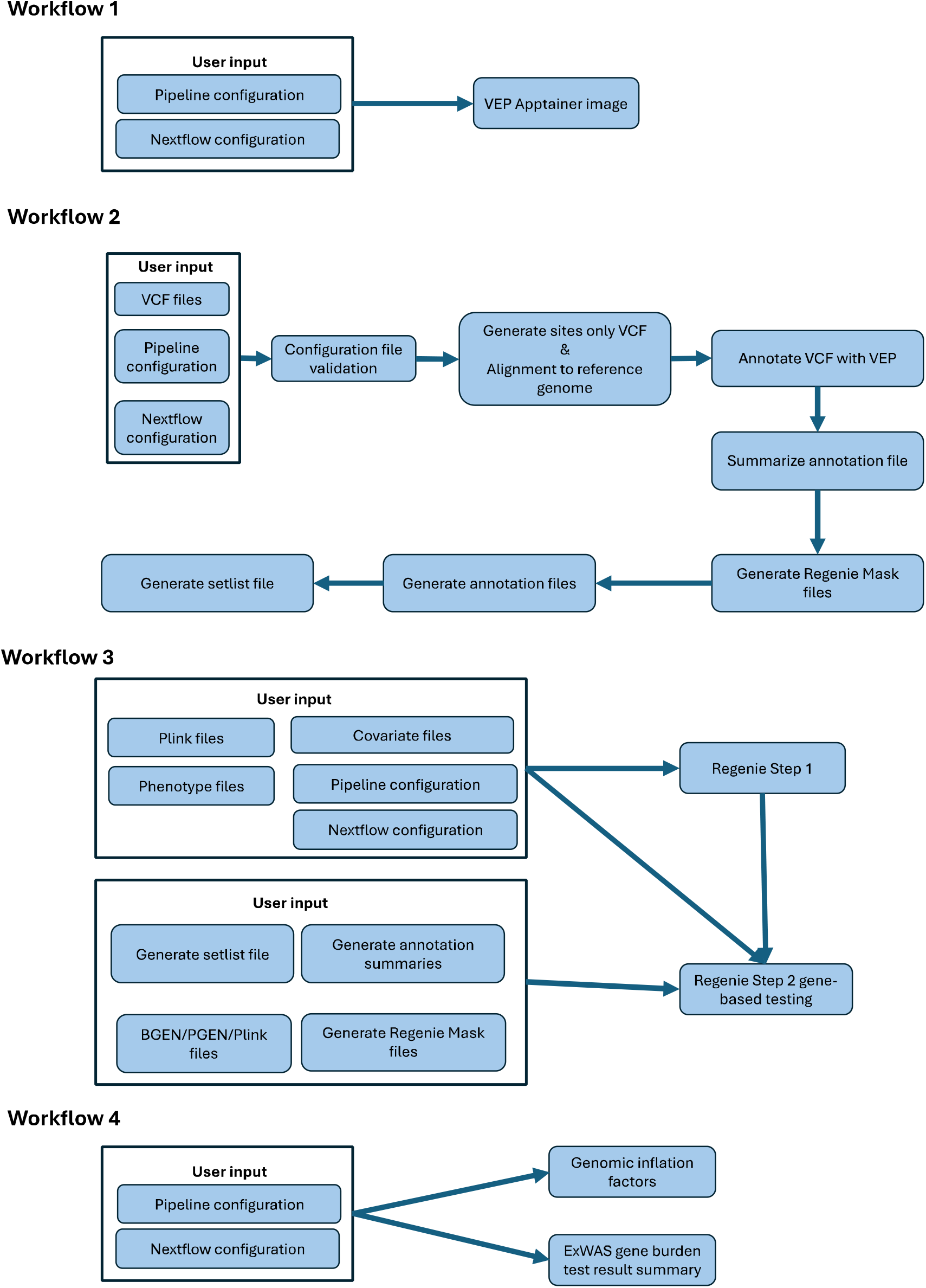
Workflow overview. All mentions of “Pipeline configuration” and “Nextflow configuration” refer the same files.

### Workflow 2: Variant annotation and Regenie input preparation

#### Workflow description and deliverables

The objective of this workflow is to annotate variants and prepare input files for Regenie. The inputs for this workflow are VCF files containing variants to be analyzed. The expected deliverables for this step include variant annotations, summaries of variant annotations, and Regenie input files as outlined in supplementary table 1.

#### Variant annotations

We used VEP 105.0 for variant annotation in this study. VEP is containerized within an Apptainer image, which is built automatically in workflow 1 (Fig 1, Table S1). The Apptainer definition file is provided on GitHub. Users are free to modify VEP version in the provided configuration file.

Sites-only VCF files are created and left-aligned to a genome assembly provided by the user to standardize allele designations. For this study, VCF files are left aligned to GRCh38 accession number GCA_000001405.15. It is recommended to keep the original variant ID; however, the user can specify in the configuration file to use chr:pos:ref:alt designation instead.

VEP plugins and corresponding cache files must be downloaded and installed by the user (https://useast.ensembl.org/info/docs/tools/vep/script/vep_plugins.html). VEP is executed in the workflow with the following parameters: *offline, symbol, stats_text, force_overwrite* and *compression_output* as bgzip. Parallel processing with VEP using the fork option is possible, though users must ensure all plugins are compatible with this option (notably, the loftee plugin does not support parallel processing (https://github.com/konradjk/loftee/issues/45).

VEP outputs are summarized as follows. The most severe prediction for each plugin is kept across all transcripts. Variant consequences (e.g., frameshift, missense, stop gained, etc), as predicted by VEP are recorded for the corresponding transcripts. Consequently, the summarized VEP results are unique for each variant, plugin, and gene combination. The results are stored in an indexed SQLite database (https://www.sqlite.org/) for efficient lookup.

There are three points to consider when using this file. Firstly, as this step is performed per plugin, variant predictions from each plugin may originate from different transcripts. Secondly, variants without gene annotations from VEP are omitted. Lastly, variants may be assigned to two or more genes with different consequences, in which case, all unique instances are kept.

#### Regenie input file preparation

Based on the summarized variant annotations, the workflow will create the mask files, the annotation file, and the setlist file based on user configuration. The format of input files created is described in the Regenie documentation (https://rgcgithub.github.io/regenie/). The gene positions in the setlist file are defined as the smallest positions across all variants assigned to the corresponding gene.

### Workflow 3: Exome-wide variant gene burden test

We used Regenie in this workflow to perform gene burden tests. A single set of genetic data in plink format is expected for Step 1 of Regenie to compute background genetic relatedness. The second step can take in pgen, bgen, or plink files. All parameters normally passed to Regenie can be specified in the configuration file.

### Workflow 4: ExWAS summary

Basic summary information is generated for each ExWAS study. A study is defined as a collection masks, gene-collapsing test models, and phenotypes. Multiple studies can be defined in the configuration and will be executed sequentially.

Two pieces of summary information are provided. Firstly, genomic inflation factors are computed for each phenotype, gene-collapsing test model, and mask. The genomic inflation factor is computed as 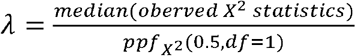, where 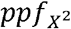 is the percent point function (or quantile function) of the X^2^ distribution. Secondly, a list of the most significant associations and the total number of exome-wide significant associations per study (i.e., across all masks, phenotypes, and gene collapsing test models) are provided. By default, a conservative Bonferroni corrected p-value threshold assuming 20,000 genes is used (i.e., the significance p-value threshold is 0.05/20,000/Number of masks/Number of phenotypes/Number of gene burden test models). This threshold is also used to determine significance status in our replication studies.

### Materials and methods for replication

For the evaluation of workflow and replication of existing studies, we used whole exome sequencing data from UK Biobank. This is the largest WES data publicly available and is one of the most used datasets for whole exome studies. Before any analyses, we performed basic quality control (QC) on all variants in the WES joint-called files. Briefly, we first selected only variants with a “PASS” filter. Then we performed genotype QC from autosomes and chromosome X separately. For autosomes, genotypes were set to unknown (./.) if they had a read depth less than 10, genotype quality less than 20, or allele balance ratio test p-value less than 0.001. For the X chromosome, the same threshold was used except for a read depth of 5 for males. Afterwards, we excluded all sites with Hardy-Weinberg mid p-value less than 5×10^−8^ or with genotype missing rate more than 10%.

### Replication study: Backman et al 2021

We evaluated nine continuous traits and compared their results to those reported by Backman and colleagues who performed ExWAS gene-burden tests using Regenie based on the UK Biobank WES data^12^ (Table S2). The nine traits from Backman et al 2021 were obtained from the GWAS catalog (Table S2). These traits were selected as they are widely studied with well-established causal genes.

### Pipeline configuration

The configuration file for replicating the Backman et al 2021 study is provided in the GitHub repository (see Data availability). Briefly, pLoF variants were defined as variants annotated as stop gained, start lost, splice donor, splice acceptor, stop lost, or frame shift variants. Only missense variants (i.e., those not classified as pLoF variants) were considered for further deleterious variant annotations by LRT, MutationTaster, Polyphen2 HDIV, Polyphen2 HVAR and SIFT. Annotation results from these tools were obtained from the dbNSFP 4.8^13^.

Two masks were used. (1) pLoF variants only or (2) pLoF variants and deleterious variants (i.e., variants annotated as deleterious by all five prediction algorithms). Each mask was tested under five minor allele frequency thresholds (singletons, 1%, 0.1%, 0.01%, and 0.001%).

### Replication study: Chen et al 2023

We next attempted to replicate the results from Chen et al 2023^14^. Chen and colleagues evaluated the value of Alphamissense in ExWAS gene burden tests across 24 common traits and diseases. In this study, we compared ExWAS gene burden test results for 11 continuous and 10 binary traits (Table S2). ExWAS results were obtained from the authors of the original publication.

### Pipeline configuration

The configuration file for replicating Chen et al 2024 is provided in the GitHub repository (see Data availability). Briefly, pLoF variants were defined as “HIGH” impact variants predicted by VEP (https://useast.ensembl.org/info/genome/variation/prediction/predicted_data.html). Deleterious variants were defined the same as above.

Two masks were used. They were (1) pLoF variants only or (2) pLoF variants and deleterious variants (i.e., variants annotated as deleterious by all five prediction algorithms). Each mask was tested under three minor allele frequency thresholds (singletons, 1%, and 0.1%).

### Replication study: Zhao et al 2024

Zhao and colleagues assessed the role of protein truncating variants in obesity using WES data from UK Biobank^4^. Different from the prior two studies, Zhao et al used variant inclusion criteria based on REVEL^15^ and performed association test using BOLT-LMM^16^.

### Pipeline configuration

The configuration file for replicating the results from Zhao et al 2024 is provided in the GitHub repository (see Data availability). Briefly, pLoF variants were defined as high-confidence loss of function variants predicted by Loftee^17^. Only missense variants were considered for deleterious variant annotation. Two overlapping missense variants inclusion criteria were used: missense variants with a REVEL score above 0.5 and missense variants with a REVEL score above 0.7.

Three masks were used: (1) pLoF only, (2) missense variants with REVEL score above 0.5, and (3) missense variants with REVEL score above 0.7. Each mask was tested under one minor allele frequency thresholds (0.1%).

## Results

### Replication study: Backman et al 2021

The correlation between p-values across all nine traits for both masks was moderate to high (Spearman correlation: 0.57 – 0.98) (Fig 2, S1). Traits with less significant results generally had lower correlations. This was the case for diastolic and systolic blood pressure. A similar pattern was observed for both masks (i.e., pLoF only, or pLoF and deleterious variants) (see Methods for mask definition details).

**Fig 2:**
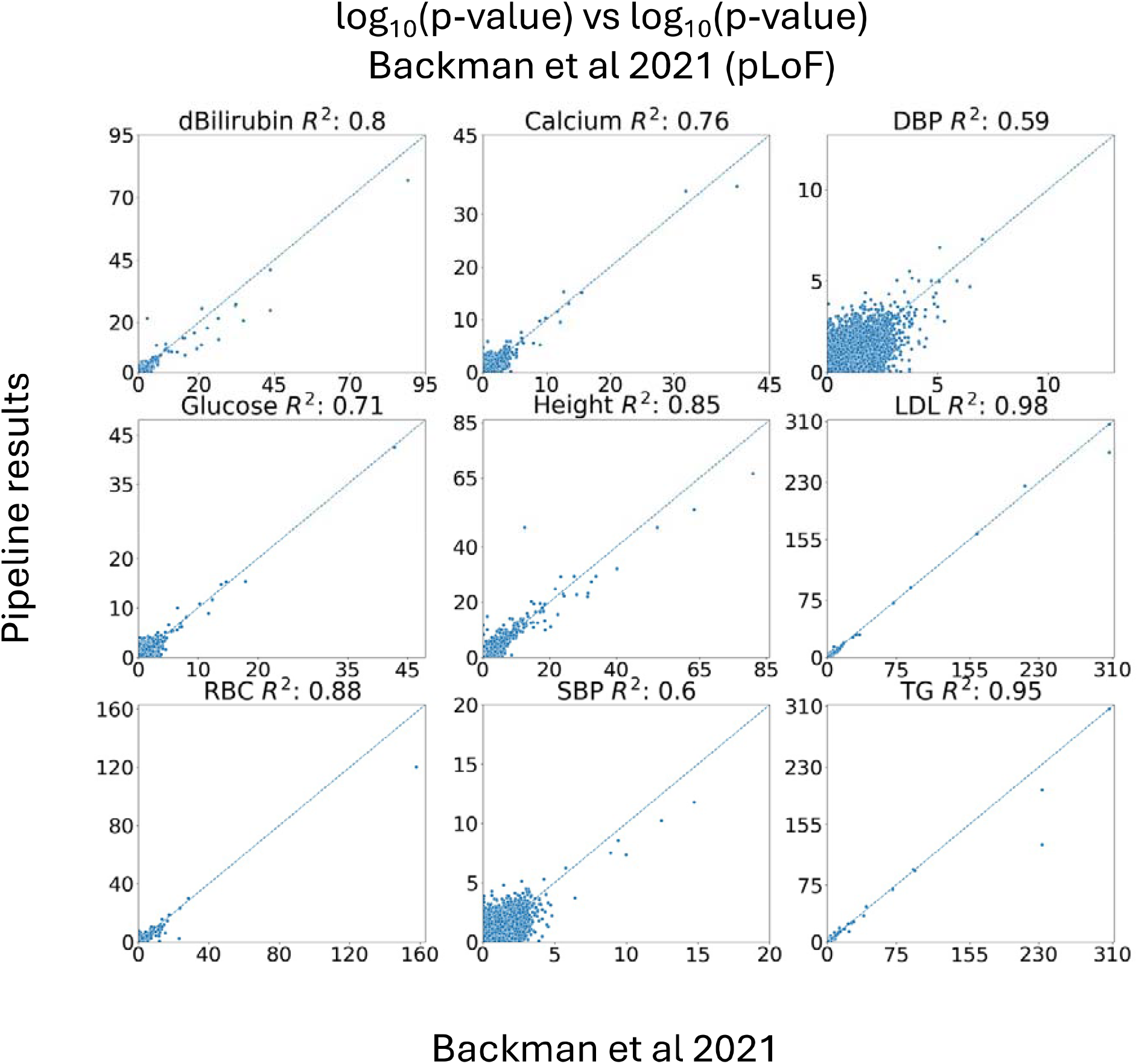
Correlation of p-values between pipeline results and those from Backman et al 2021 based on pLoF variants. P-values were negative log 10 transformed. Pearson correlations are reported.

Based on a conservative Bonferroni correction threshold of 2.5 × 10^−7^ (assuming 20,000 genes, across 2 masks, 5 allele frequencies threshold, and 9 phenotypes), with the exception of DBP, 72% to 100% of significant results were replicated (Table S3). DBP yielded only five significant results with the two most robust findings replicated by our pipeline. The low replication rate for DBP aligns with the fact that DBP had the weakest p-value correlation among the nine phenotypes (0.57–0.59) and generally had less significant results (Fig 2, S1). Overall, for the results that did not replicate, the p-values in our replication were generally near the Bonferroni-adjusted p-value threshold (Fig S5).

### Replication study: Chen et al 2023

The correlations across continuous and binary traits were moderate to high (0.79 – 0.99) and likewise, no differences were seen between masks (Fig 3, S2, S3, S4). In general, the correlations of p-values among continuous traits were slightly stronger than among binary traits. Like before, the correlations were the strongest for traits with strong significant results (e.g., triglyceride and low-density cholesterol) and weaker for traits with less significant results (e.g., diastolic and systolic blood pressure).

**Fig 3:**
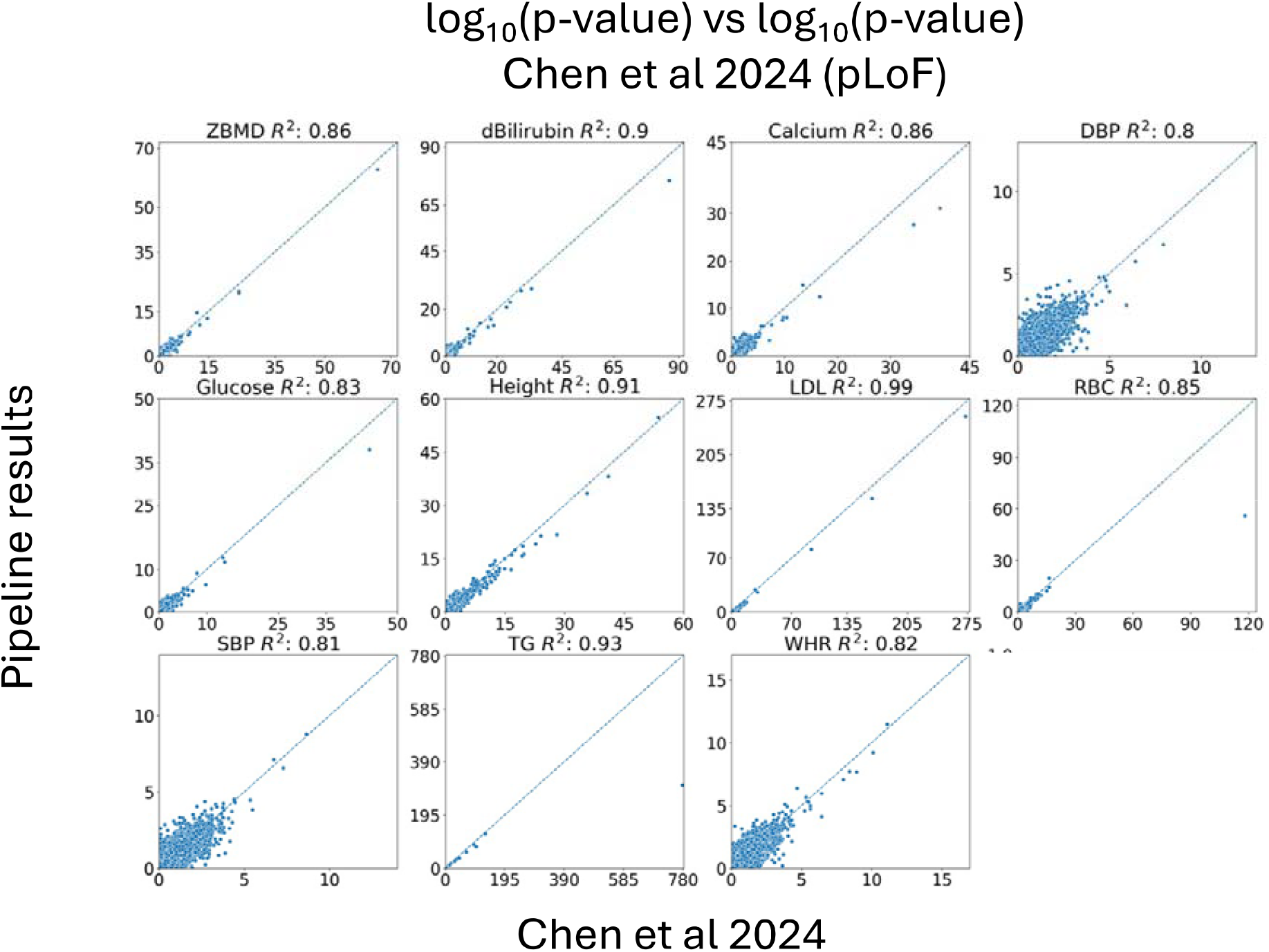
Correlation of p-values between pipeline results and those from Chen et al 2024 based on pLoF variants for 11 continuous traits. P-values were negative log 10 transformed. Pearson correlations are reported.

Anywhere from 50% to 100% of the gene-level significant results identified by Chen et al based on a conservative Bonferroni threshold of 1.98 × 10^−8^, assuming 20,000 genes for 2 masks, 3 allele frequencies and 21 phenotypes, were replicated. The lowest replication rate was for hypertension for which there were 2 significant hits identified by Chen et al of which 1 was replicated (Table S3). Likewise, for genes that did not replicate, the p-values in our replication were generally close to the Bonferroni-adjusted threshold (Fig S6).

We also compared annotation results between those generated by the pipeline and those generated by Chen et al. There was good overlap between both sets of annotations where 99% of pLoF variants (841,113/847,306) and 98% of missense deleterious variants (1,092,410/1,114,795) were shared (Fig 4). Importantly, the pLoF variants annotated by the pipeline and the missense deleterious variants identified by Chen et al., and the deleterious variants annotated by the pipeline and the pLoF variants identified by Chen et al., were mutually exclusive.

**Fig 4:**
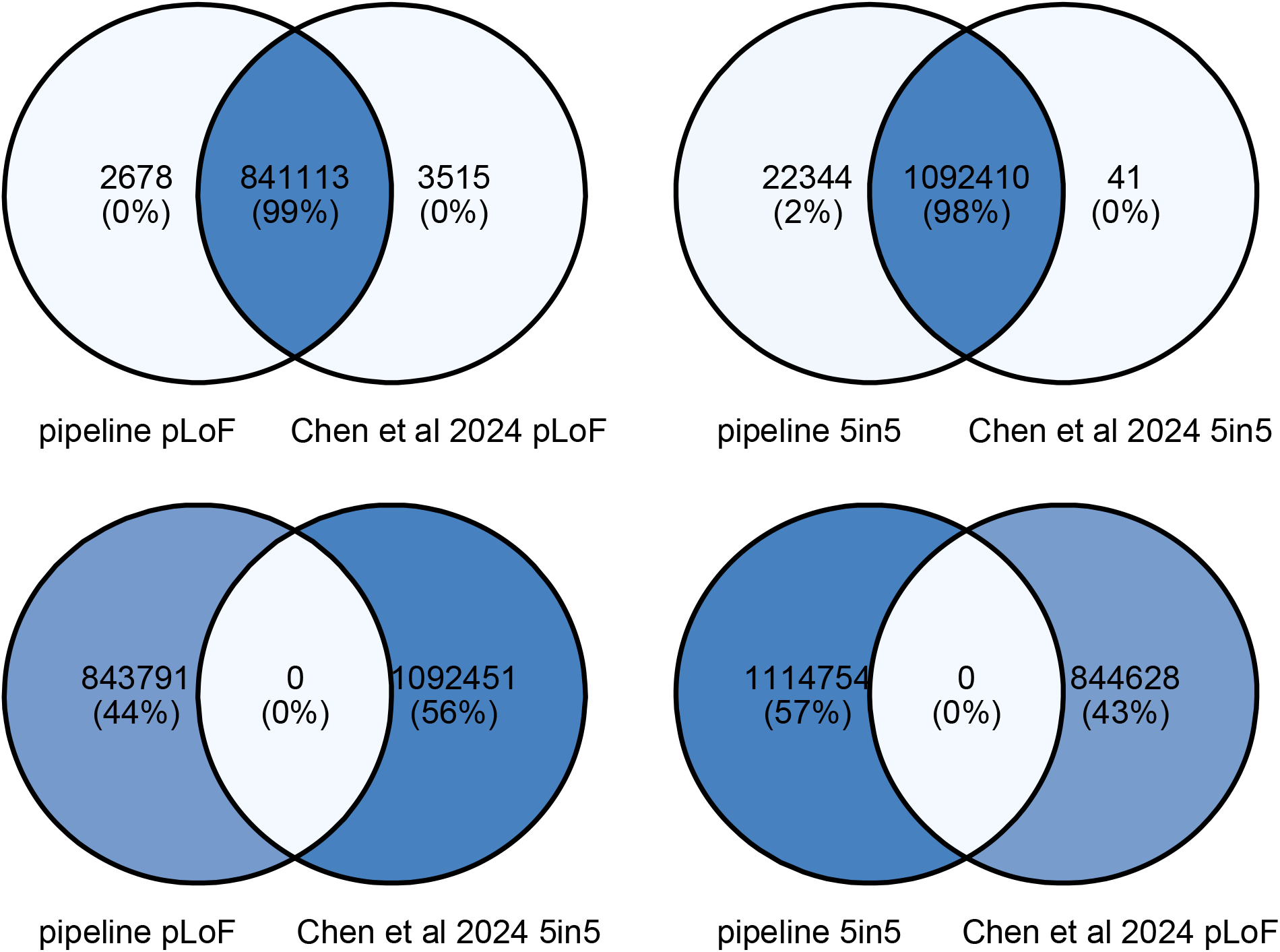
Comparison of variant annotations between pipeline results and those from Chen et al 2024.

The difference in annotation was due to differences in the VEP version used. In total, the pipeline identified 843,791 pLoF variants and Chen et al identified 844,628 pLoF variants. 3,515 pLoF variants were uniquely identified by Chen et al. Across the 3,515 pLoF variants uniquely identified by Chen et al, but not by the pipeline, 3,512 had lower impact annotations (i.e., low, moderate, or modifier), and hence were not annotated as pLoF. The three remaining variants were assigned to intergenic regions; hence were not included in the pipeline. The 41 deleterious variants uniquely identified by Chen et al did not have complete annotation results from all five prediction algorithms. Despite this difference, it is worth noting that ExWAS gene burden test results were largely consistent.

### Replication study: Zhao et al 2024

In a recent publication, Zhao and colleagues identified protein truncating variants in the *BSN* gene with much larger effects on BMI than previously known genes such as *MC4R*^4^. They performed exome-wide gene-burden testing using whole exome sequencing (WES) data from the UK Biobank project among others.

ExWAS gene-based testing results for the nine significant genes identified by Zhao et al were obtained from supplementary table 1 of the original publication. Within our dataset, the *TOX4* gene had 2 missense variants with a REVEL score above 0.7. Consequently, it was dropped from the gene-burden association analysis. The correlation between negative log10 transformed p-value was strong (Pearson correlation R^2^ = 0.99) (Fig 5A). Similarly, the effect sizes were replicated with overlapping confidence intervals (Fig 5B).

**Fig 5:**
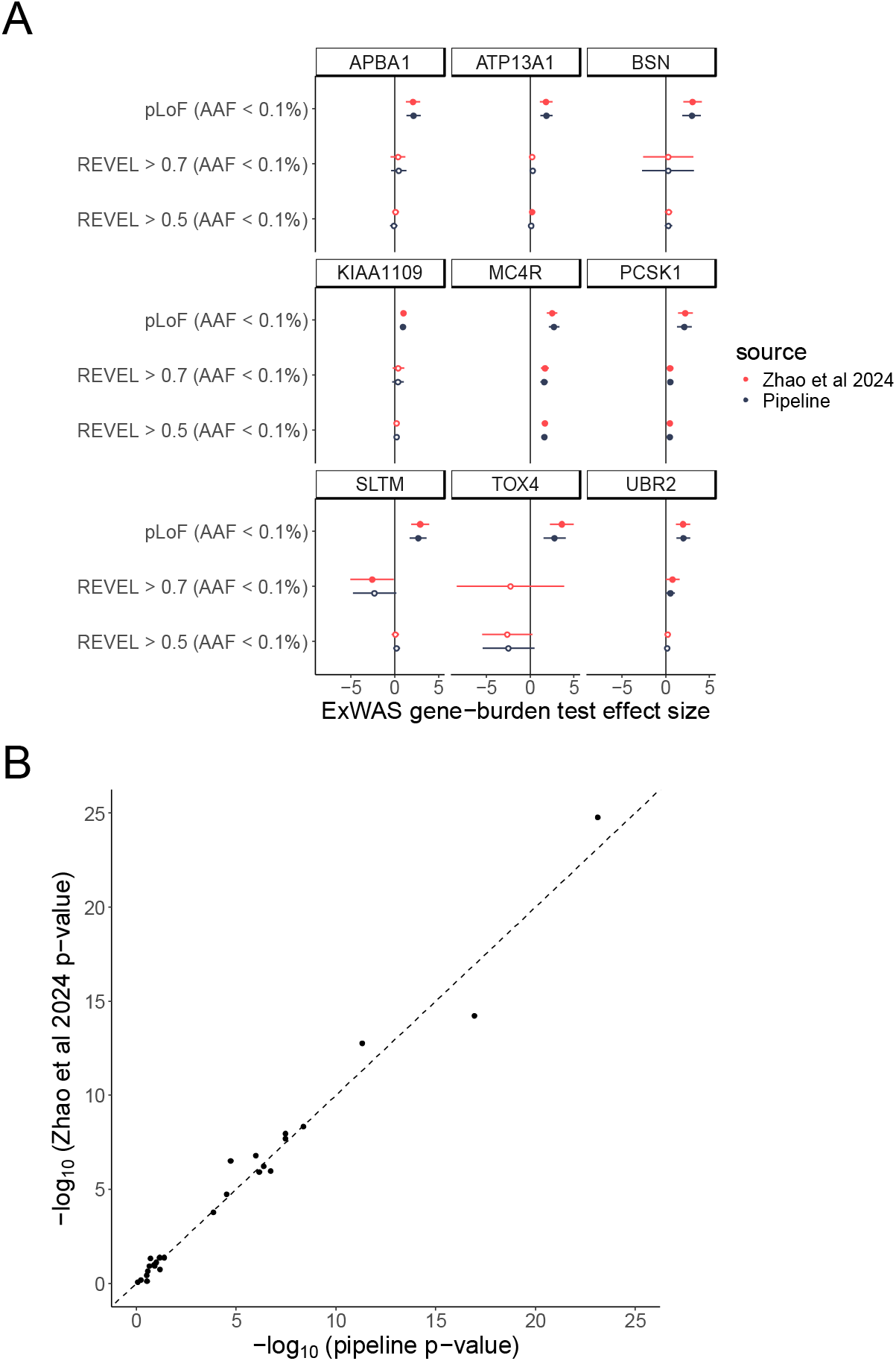
Comparison of results produced by the pipeline and those obtained from Zhao et al 2024. A) The effect size of the 9 genes reported by the authors together with the 95% confidence interval is shown. The *TOX4* gene only had 2 variants with Revel score above 0.7 and minor allele frequencies less than 0.1%, thus were excluded in the pipeline analysis. B) Correlation of p-values between the two studies.

## Discussion

In this study, we provided a standardized flexible workflow for performing exome-wide association gene-burden testing using well-established bioinformatics tools. Starting from standard genetic data, this workflow performs variant annotations, ExWAS gene burden tests, and produces a basic summary of results. We applied this workflow to replicate three recent ExWAS publications. The configuration files for replicating each study using the workflow are provided in the GitHub repository. This tool is actively maintained, with regular updates and incorporation of new analytical workflows.

## Data Availability

The configuration files for each replication study are provided on the GitHub repository (https://github.com/richardslab/EXWAS_pipeline). ExWAS results from Backman et al, Zhao et al, and Chen et al can be obtained from GWAS catalog or the original publication.

## Disclosure

JBR has served as an advisor to GlaxoSmithKline and Deerfield Capital. JBR is the CEO of 5 Prime Sciences. YC is an employee of 5 Prime Sciences. No conflict of interests is declared for the other authors.

## Funding statement

The Richards research group is supported by the Canadian Institutes of Health Research (CIHR: 365825; 409511, 100558), the Lady Davis Institute of the Jewish General Hospital, the Canadian Foundation for Innovation, the NIH Foundation, Cancer Research UK, Genome Québec, the Public Health Agency of Canada, Genome Québec, McGill University and the Fonds de Recherche Québec Santé (FRQS). JBR is supported by a FRQS Clinical Research Scholarship. Support from Calcul Québec and Compute Canada is acknowledged. This work was supported by Cancer Research UK [grant umber C18281/A29019]. TwinsUK is funded by the Welcome Trust, Medical Research Council, European Union, the National Institute for Health Research (NIHR)-funded BioResource, Clinical Research Facility and Biomedical Research Centre based at Guy’s and St Thomas’ NHS Foundation Trust in partnership with King’s College London. KYHL was supported by CIHR doctoral scholarship. These funding agencies had no role in the design, implementation or interpretation of this study.

## Acknowledgement

ChatGPT (OpenAI) and Grammarly (Grammarly Inc) were used to check grammar and improve writing style for clarity and conciseness. UK Biobank was conducted under the project number 27449.

