## Supplementary Figures for "A flexible pipeline for reproducible exome-wide rare variant gene-trait associations"

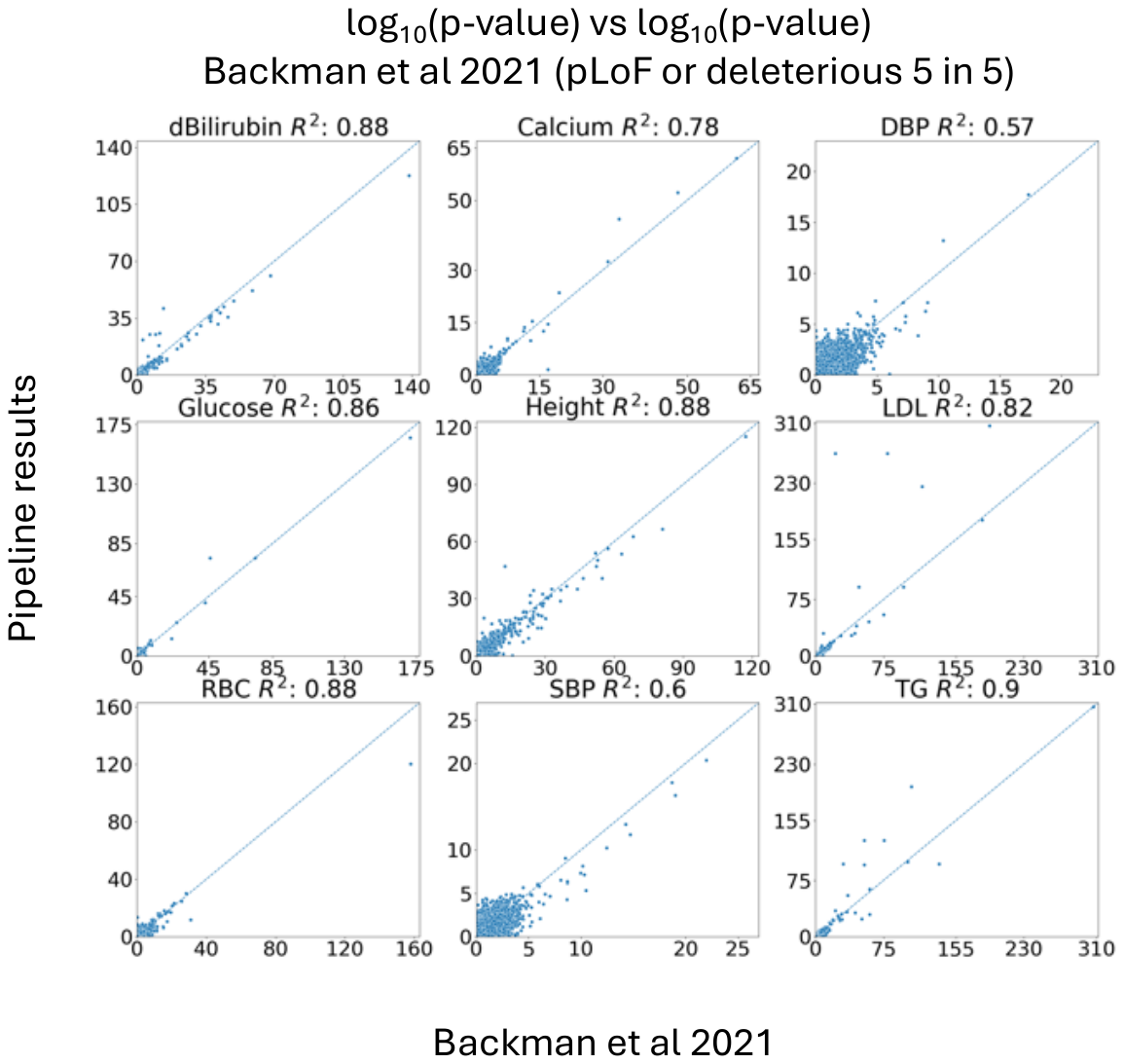

Fig S1: Correlation of p-values between pipeline results and those from Backman et al 2021 based on pLoF variants and deleterious variants (i.e., those annotated as deleterious by all five prediction algorithms). P-values were negative log 10 transformed. Pearson correlations are reported.

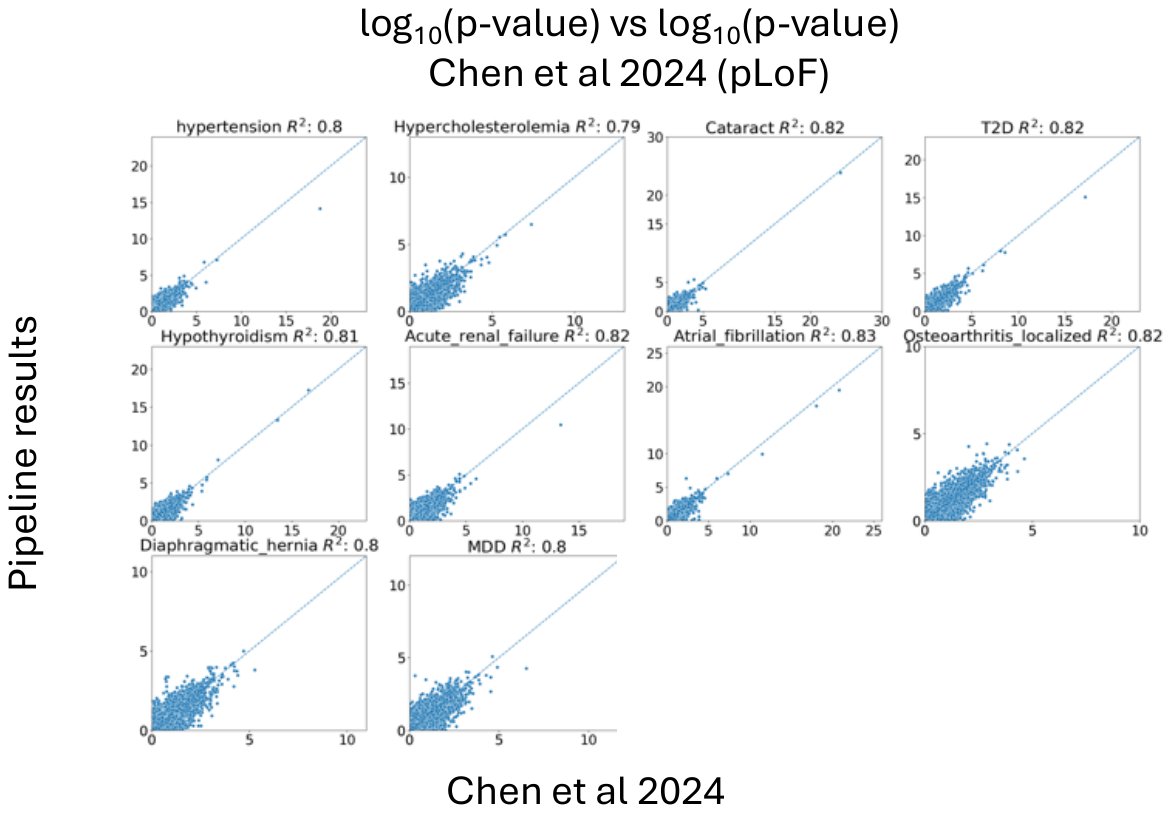

Fig S2: Correlation of p-values between pipeline results and those from Chen et al 2024 based on pLoF variants for 10 binary traits. P-values were negative log 10 transformed. Pearson correlations are reported.

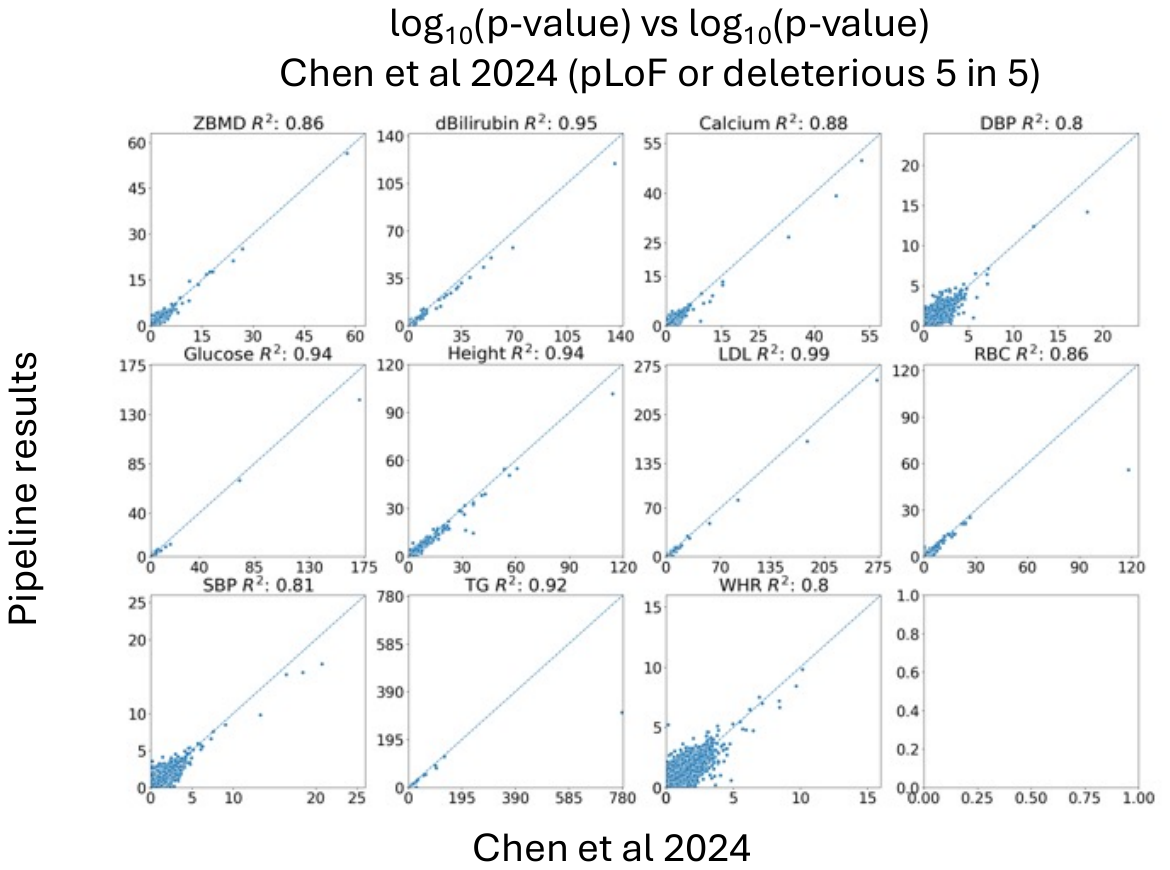

Fig S3: Correlation of p-values between pipeline results and those from Chen et al 2024 based on pLoF variants and deleterious variants (i.e., those annotated as deleterious by all five prediction algorithms) for 11 continuous traits. P-values were negative log 10 transformed. Pearson correlations are reported.

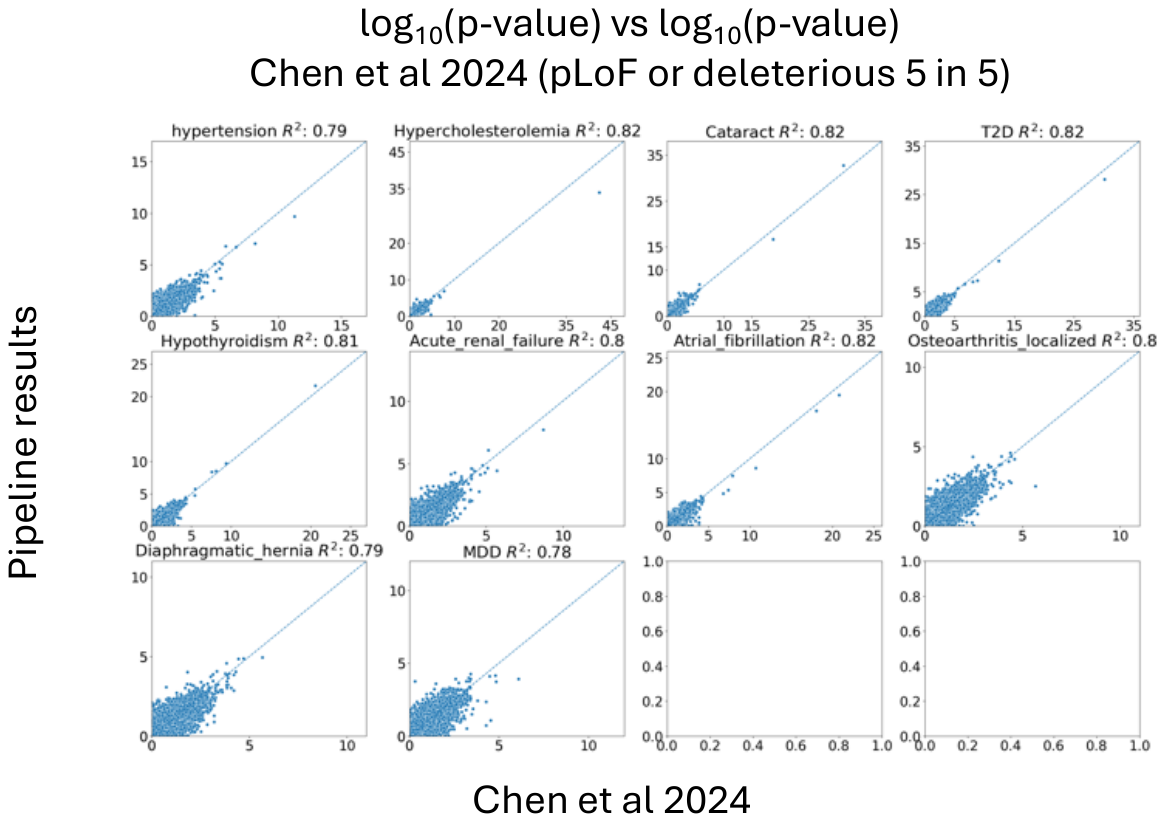

Fig S4: Correlation of p-values between pipeline results and those from Chen et al 2024 based on pLoF variants and deleterious variants (i.e., those annotated as deleterious by all five prediction algorithms) for 10 binary traits. P-values were negative log 10 transformed. Pearson correlations are reported.

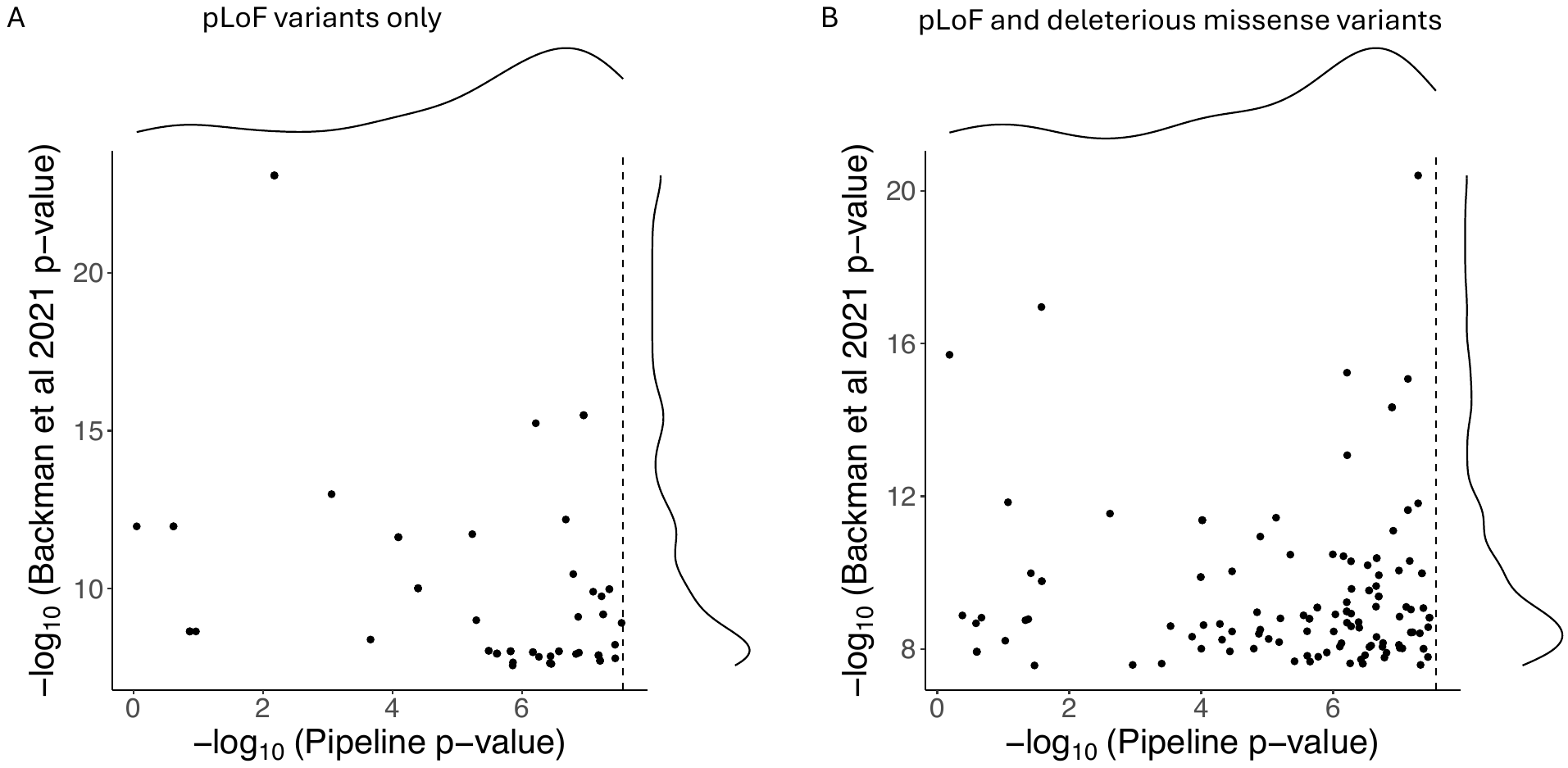

Fig S5: Comparison of p-values among genes with significant gene burden test results from Backman et al 2021 but insignificant results from the pipeline. Dotted line indicates Bonferroni corrected significance threshold accounting for 2 variant masks, 5 allele frequency thresholds, 20,000 genes and 1 gene burden testing models.

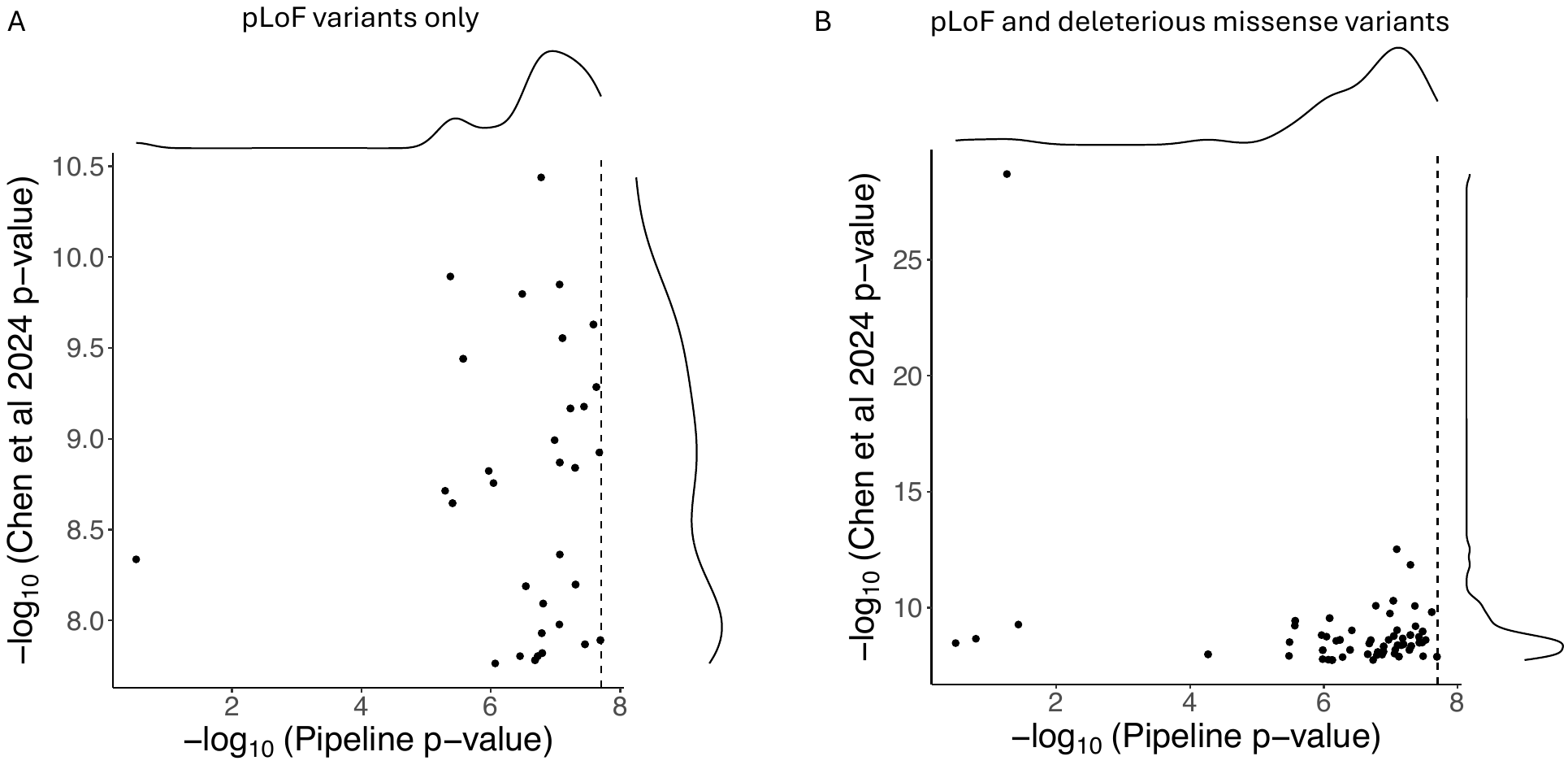

Fig S6: Comparison of p-values among genes with significant gene burden test results from Chen et al 2024 but insignificant results from the pipeline. Dotted line indicates the Bonferroni corrected significance threshold accounting for 2 variant masks, 3 allele frequency thresholds, 1 burden test models and 20,000 genes
